# Total Added Sugar Consumption is not Significantly Associated with Risk for Prediabetes Among U.S. adults: National Health and Nutrition Examination Survey, 2013-2018

**DOI:** 10.1101/2022.08.22.22279065

**Authors:** Nadia Markie Sneed, Andres Azuero, Jacqueline Moss, Amy M. Goss, Shannon A Morrison

## Abstract

**Background:** Prediabetes affects 34.1% U.S. adults and is primarily linked to added sugars consumed from sugar-sweetened beverages. It is unclear if total dietary intake of added sugar also increases the risk for prediabetes.

**Objective:** This study examined if total (g/day) and percent intakes of <10%, 10-15%, or >15% added sugar increase the odds for prediabetes in U.S. adults.

**Design:** A cross-sectional, secondary analysis using 2013-2018 NHANES data was conducted.

**Participants/setting:** This study included data from U.S. adults ≥ 20 years with normoglycemia (N= 2,154) and prediabetes (N= 3,152) with 1-2 days of dietary recall information.

**Main outcome measures:** Prediabetes, defined as a Hemoglobin A1c of 5.7%-6.4% or a fasting plasma glucose of 100-125 mg/dL.

**Statistical analysis:** Survey-weighted logistic regression was used to estimate odds ratios of prediabetes based on usual intakes of added sugar (total and percent intakes) using the National Cancer Institute Method. Differences in prediabetes risk and total and percent intakes of added sugar were compared by race/ethnicity.

**Results:** The sample’s total energy intake from added sugar was 13.9%. Total (unadjusted: OR: 1.01, 95% CI: .99 - 1.00, *p* = .26; adjusted: OR: 1.00, 95% CI: .99 - 1.00, *p* = .91) and percent intakes of added sugar (unadjusted [<10%: (ref); 10-15%: OR: .93, 95% CI: .77 - 1.12, *p* = .44; >15%: OR: 1.03, 95% CI: .82 - 1.28, *p* = .82] and adjusted [<10%: (ref); 10 - 15%: OR: .82, 95% CI: .65 - 1.04, *p* = .09; >15%: OR: .96, 95% CI: .74 - 1.24, *p* = .73]) were not significantly associated with an increased odds of prediabetes. Prediabetes risk did not differ by race/ethnicity for total (unadjusted model [*p* = .65]; adjusted model [*p* = .51]) or percent (unadjusted model [*p* = .21]; adjusted model [*p* = .11]) added sugar intakes.

**Conclusions:** In adults ≥20 years with normoglycemia and prediabetes, total added sugar consumption did not significantly increase one’s risk for prediabetes and risk estimates did not differ by race/ethnicity. Experimental studies should expand upon this work to confirm these findings.

## Introduction

Prediabetes is a relatively asymptomatic, but serious medical condition characterized by insulin resistance and intermittent hyperglycemia^1^ that affects approximately 88 million U.S. adults.^2^ Prediabetes is a precursor to type 2 diabetes (T2D) ^1^ and is associated with chronic kidney disease^3^ and cardiovascular disease risks (i.e., hypertension, dyslipidemia),^4^ independent of T2D progression.^3,4^ Significant disparities in the prevalence of prediabetes are observed among minority populations, particularly for non-Hispanic Black and Hispanic adults, in comparison to non-Hispanic White adults (32%, 35.3%, 31%, respectively).^5^ Due to the slow but progressive nature of prediabetes pathology, roughly 85% of adults are unaware of their condition^2,6^ and often remain unaware until after the condition has progressed to T2D.^7^ Diabetes is considered a preventable diet-related disease,^8^ yet, about 5-10% of adults with prediabetes progress to T2D annually and 70% of adults with prediabetes develop T2D within their lifetime.^7,9^ Whether factors such as diet, genetics, advancing age, other lifestyle choices (e.g., physical activity) or a combination of these factors increase the risk for prediabetes is not fully known. However, longitudinal, observational studies examining the role of nutrition on metabolic conditions suggest diet is a primary predictor of a plethora of cardiac and metabolic health conditions, including prediabetes.^10-14^

In the 1950s and 1960s cardiovascular disease was hypothesized to be a consequence of excessive fat consumption and subsequently hyperlipidemia.^15^ Thus, a change in the recommended intake of dietary fats and carbohydrates occurred in the late 1970s^16^ and the first published U.S. Dietary Guidelines recommended decreasing dietary fat from 40% to 30% and increasing carbohydrate intake to approximately 55% - 60% of total daily energy intake.^15^ Unfortunately, this shift paralleled a drastic rise in obesity and diabetes rates across the 1980s and 1990s and had little influence on cardiovascular disease prevalence.^2,17^ Concurrent with the shift in diet trends, food manufacturers increased production of carbohydrate rich, low-fat foods. In short, food manufacturers substituted carbohydrates in lieu of fats, largely in the form of added sugars across a multitude of foods and beverages,^18^ and these ultra-processed, sugary foods became a mainstay in the U.S. diet.^19^

Added sugars are caloric sweeteners added to foods and beverages during processing, preparation, or prior to consumption^18^ and the most common types are sucrose, used predominately in solid foods, and high-fructose corn (HFCS), used predominately in sugar-sweetened beverages (SSBs).^20^ Evidence linking obesity and metabolic disease to added sugar prompted an additional modification to the U.S. Dietary Guidelines.^21^ In 2015, total dietary intake of added sugar was recommended to not exceed 10% of an individual’s daily caloric intake,^21^ a recommendation that persists today.^22^ Nonetheless, an average of 270 calories (more than 13% total energy intake) from added sugar is consumed by U.S. adults daily and exacerbates the issue of overconsumption.^22^

Both natural sugars (found in fruits/vegetables) and added sugars contain the monosaccharide sugars glucose and fructose.^18^ Yet, evidence suggests that chronic consumption of high added sugar diets (i.e., ∼15-25% total energy intake)^23^ containing fructose are primarily responsible for T2D risk^24,25^ and are independent of total energy intake or body mass index (BMI).^25-28^ Directly, fructose metabolism increases hepatic lipid synthesis (i.e., de novo lipogenesis) and promotes a reduction in hepatic fatty acid oxidation resulting in fatty liver and subsequent hepatic insulin resistance.^20,29^ The correlation between added sugar and prediabetes is strongly linked to dysregulated fructose metabolism,^30^ yet studies assessing the direct effects of added sugar on prediabetes have been limited with most examining added sugar proxies such as SSBs, HFCS, and fructose-sweetened beverages and not total added sugar from all dietary sources.^25,31-40^ Thus, the potential mechanisms and metabolic effects of total added sugar consumption on prediabetes risk are still under debate by scientists and warrant further investigation.^23^

Also of concern is that minority populations demonstrate significant health disparities in obesity and T2D prevalence in comparison to non-Hispanic White individuals.^5,41^ Moreover, consumption of a high carbohydrate diet in minority populations (i.e., non-Hispanic Black adults) has been shown to promote an exaggerated insulin response that occurs independent of overweight/obesity status.^42^ Non-Hispanic Black adults consume the greatest quantities of added sugar followed by non-Hispanic White adults, Hispanic adults, and non-Hispanic Asian adults (19 teaspoons (tsp), 17 tsp, 16 tsp, and 10 tsp respectively)^43^ raising the question as to whether differences in dietary intake from added sugar may contribute to these health disparities.

No studies have examined if *total* dietary intake of added sugar increases the risk for prediabetes and if so, how much (e.g., >15% total caloric intake) may be responsible for the increased risk observed. Also, it is unclear whether added sugar uniquely influences the risk for prediabetes by race/ethnicity, particularly among those that consume high quantities of added sugar (i.e., non-Hispanic Black adults).

The main objective of this study was to examine whether total added sugar consumption was associated with prediabetes in a large nationally representative sample of U.S. adults ≥20 years. Second, this study examined if total added sugar consumption, as a percentage of total energy intake (<10%, 10-15%, >15%), was associated with differing risk probabilities for prediabetes. Last, this study assessed if the associations between total and percent intakes of added sugar and prediabetes risk differed by race/ethnicity. The study’s guiding a priori hypothesis was that positive associations between total and percent added sugar intakes and prediabetes risk would be observed, including greater risk at higher added sugar intakes (e.g., >15% total energy), and that significant risk differences would be observed by race/ethnicity with increased risk among high T2D risk groups (e.g., non-Hispanic Black, Hispanic adults).

## Materials and Methods

### Study Design

This study used data from the National Health and Nutrition Examination Survey (NHANES) which is a repeated cross-sectional survey that employs a complex, multistage, probability sampling design to collect health and nutrition information from ∼5,000 noninstitutionalized U.S. civilians (age 0 years and older) annually.^44,45^ NHANES is supported by the National Center for Health Statistics (NCHS) and Centers for Disease Control and Prevention.^46^ Specific details about the design and operations of NHANES, including sampling and data collection procedures, have been previously described elsewhere.^45,47^ A cross-sectional analysis was conducted and included data collected from NHANES respondents ≥20 years during the 2013-2014, 2015-2016, and 2017-2018 NHANES cycles. NHANES study protocols are approved by the NCHS Research Ethnics Review Board^48^ and are compliant with the Health and Human Services Policy for Protection of Human Research Subjects (45 CFR part 46).^45,49^ Only de-identified, publicly-available data were analyzed, therefore the study was designated as ‘Not Human Subjects Research’ by the University of Alabama at Birmingham.^50^

### Analytic Sample

The analytic sample included respondents ≥20 years of age with fasting plasma glucose (FPG) or Hemoglobin A1c (HbA1c) defined prediabetes or normoglycemia. Respondents represented the following NHANES racial and Hispanic origin groups: Hispanic (including Mexican American and other Latino populations), non-Hispanic Black, non-Hispanic White, Asian American, and Other Race which included persons not self-identifying with any of the prior categories. The initial weighted sample (taken from the fasting subsample)^47^ included a total of 5,888 respondents ≥20 years of age that excluded pregnant and lactating women (n=115) and those taking insulin or diabetic medications (n=963). An additional 167 respondents with T2D having either a HbA1c ≥ 6.5% (48 mmol/mol) or a FPG ≥ 126 mg/dL (7.0 mmol/L) were excluded resulting in 5,721 adults. Next, 415 adults were excluded due to not having at least 1 day of dietary recall information that contained a value for added sugar. The final sample included 5,306 adults with 3,152 respondents identified as having prediabetes (HbA1c 5.7% - 6.4% [39-47 mmol/mol] and FPG 100-125 mg/dL [5.6-6.9 mmol/L]) and 2,154 identified as having normoglycemia (HbA1c <5.7% [<39 mmol/mol] and FPG <100 mg/dL [<5.6 mmol/L]).^1^

### Prediabetes Assessment

The outcome variable for this study was prediabetes which has been previously defined and was based on the American Diabetes Association “Standards of Medical Care in Diabetes - 2021” classification.^1^ As part of the NHANES data collection process, physical measurements and laboratory tests are collected during the mobile examination center visit from participants ages ≥12 years.^45^ Data files of whole blood specimens of glycohemoglobin (i.e., HbA1c) and FPG were included for analysis in this study to define prediabetes.^51,52^ NHANES collects samples at the examination center using a Tosoh G8 Automated Glycohemoglobin (HbA1c) and Cobas c311 Analyzer (FPG). Detailed NHANES laboratory procedures are reported elsewhere.^51,52^

### Dietary Intake Assessment

Dietary intake data, including added sugars and total calories, were collected for the dietary assessment component of NHANES which uses the 24-hour dietary recall method.^53^ Diet recalls are pre-announced and performed by trained interviewers using the validated U.S. Department of Agriculture’s Automated Multiple-Pass Method (AMPM) previously described elsewhere.^45,53^ The first diet recall is administered in-person during the mobile examination center visit (on either weekdays or weekends) and the second is administered over the phone 3-10 days later.^45^

Added sugars are defined as sugars, syrups, fruit juice concentrates, or caloric sweeteners added during processing, preparation, or prior to food and beverage consumption that exclude natural sugars present in dairy and fruit (including whole fruit and 100% fruit juice).^54^ Estimates for added sugar were obtained from the Food Patterns Equivalents Database (FPED) of the Food and Nutrient Database for Dietary Studies.^55^ Added sugars were reported in teaspoon equivalents consumed per subject, per day calculated from foods/beverages.^54^ Total calories from day 1 and day 2 dietary recalls were obtained from the NHANES nutrient intake files and were reported in kilocalories (kcals). The FPED files were merged with NHANES total nutrient intake files to combine estimates for added sugar and total calories. For this study, to reflect updates to nutrition facts labeling,^56^ added sugar was converted from teaspoon equivalents to grams (1 teaspoon equivalent = 4.2 grams) and from grams to calories (1 gram = 4 kilocalories) for day 1 and day 2 dietary recalls before the final dataset merge.^56,57^ Once all 2013-2018 data files were merged, three percent intake groups for added sugar (<10%, 10-15%, and >15%) were calculated by dividing grams of added sugar by total calories. The groups represent dietary guideline recommendations (<10%), average U.S. intake (∼13%), and above average intake (>15%) respectively.^22^

### Covariates

Regression models included the following covariate categories: age, gender, race/ethnicity, BMI (kg/m^2^), usual intake for total calories (kcals), physical activity, smoking status, educational attainment, and income.

Demographic variables collected during the NHANES in-home interview included age, gender, and race/ethnicity which were self-report.^58,59^ Age was reported in years and calculated using participant’s dates of birth. Participants ≥80 years were coded as ‘80’ to prevent risk of participant disclosure. Gender was classified as either “male” or “female”.^58^ Race and Hispanic origin was categorized into non-Hispanic White, non-Hispanic Black, Hispanic (including Mexican Americans and Latinos), Asian Americans, or Other Race (including persons not identifying with the previously reported categories).^47,59^ BMI was categorized using the following CDC classifications for adults: underweight (18.5 kg/m^2^), healthy weight (18.5-24.9 kg/m2), overweight (25-29.9 kg/m^2^), or obese (≥ 30 kg/m^2^).^60^ Health behaviors and sociodemographic factors (physical activity, smoking status, education, and ratio of family income to poverty) were based on self-reported questionnaire data. Physical activity was classified using the NHANES physical activity questionnaire (PAQ650 and PAQ665) and was defined as (yes/no) engagement in ≥10 minutes of moderate and/or vigorous recreational activity during a typical week.^61^ Smoking status was defined as either current smoker (tobacco use within the last 5 days) or non-smoker (no reported use within last 5 days).^62^ Education level was defined as having either less than a high school degree, having a high school degree or GED, or having more than a high school degree.^59^ Ratio of family income to poverty was taken from the NHANES demographic questionnaire and was categorized using the family monthly poverty level index categories calculated by NHANES (≤1.30, >1.30 to 1.85, and >1.85) which represents common poverty guideline percentages.^63^

### Statistical Analysis

All analyses were performed using SAS Studio version 3.8, Enterprise Edition.^64^ NHANES analytic guidelines^47^ were followed using SAS procedures^65^ appropriate for complex survey designs. Survey data from 2013-2014, 2015-2016, and 2017-2018 were combined and appropriate sampling weights (from the fasting subsample WTSAF2YR) were created for the combined dataset and applied to all models prior to analyses to account for differential nonresponse and planned oversampling of certain subgroups.^47^ Data on characteristics were reported using means and standard errors for continuous variables and percentages and standard errors for categorical variables. Characteristics were reported for the overall sample and by normoglycemia or prediabetes status. Rao Scott chi square tests were used to examine differences in sample characteristics for categorical variables by normoglycemia and prediabetes status. Ordinary least squares regression was used to examine differences in sample characteristics for continuous variables by normoglycemia and prediabetes status.

The National Cancer Institute (NCI) method^66,67^ was used to estimate usual intake of added sugar and total caloric intake using day one and day two 24-hour dietary recall data. The NCI method requires two or more dietary recalls on nonconsecutive days for a random subset of the population to account for between- and within-person variation in intake and can be used to correct for measurement error when estimating usual intake of nutrients.^66^ A 2-step process was used to estimate usual intake of added sugar and total daily calories using the MIXTRAN and INDIVIT macros provided by the NCI.^66,68^ In step 1, the MIXTRAN macro was used to generate an “amount-only” model of daily consumed nutrients (i.e., added sugar) using 24 hour dietary recall data on a transformed scale.^68^ Intake day of the week^69^ was included as a covariate in the MIXTRAN model to account for possible weekday or weekend day effects on dietary intake.^70^ In step 2, INDIVINT was used to estimate usual intake for total calories and added sugar with parameters estimated from step 1. The INDIVINT macro was selected because it can be used to predict individual nutrient intakes for use as predictors in a disease model.^66,68^

Survey weighted logistic regression was used to test whether usual intake of total and percent added sugar intakes were associated with an increased odds of prediabetes relative to normoglycemia. Usual intake of added sugar was modeled as a continuous variable (g/day) and non-linear associations for added sugar as a percentage of total energy intake were examined (<10%, 10-15%, >15% kcal/day). A dichotomous indicator for prediabetes was constructed from HbA1c and FPG values. To aid in interpretation, estimated risks for prediabetes by total added sugar (g/day) were reported for mean and tertial intakes. Additionally, estimated risks for prediabetes by percent intakes of added sugar were reported as <10%, 10-15%, and >15% total energy from added sugar in kcals/day. Adjusted models included the following covariates: age in years, gender, race/ethnicity, BMI (kg/m^2^), total energy intake (kcal/day), engagement in physical activity, smoking status, education level, and PIR. Interaction terms between added sugar and race/ethnicity were used to examine differences in the relationship between prediabetes risk and total and percent added sugar intakes by race/ethnicity for non-Hispanic White, non-Hispanic Black, Hispanic, Asian American and Other Race respondents. All tests were two-sided and a *p* value <.05 was considered statistically significant.

## Results

### Sample Characteristics

A total of 5,306 adults with normoglycemia (41% of the sample) and prediabetes (59%) were included in the weighted sample and reported consuming 13.9% of their total daily calories from added sugar. There were no statistically significant differences in consumption of added sugar between groups (normoglycemia vs. prediabetes). Table 1 shows the overall characteristics of adults ≥20 years and by normoglycemia and prediabetes status.

**Table 1.**
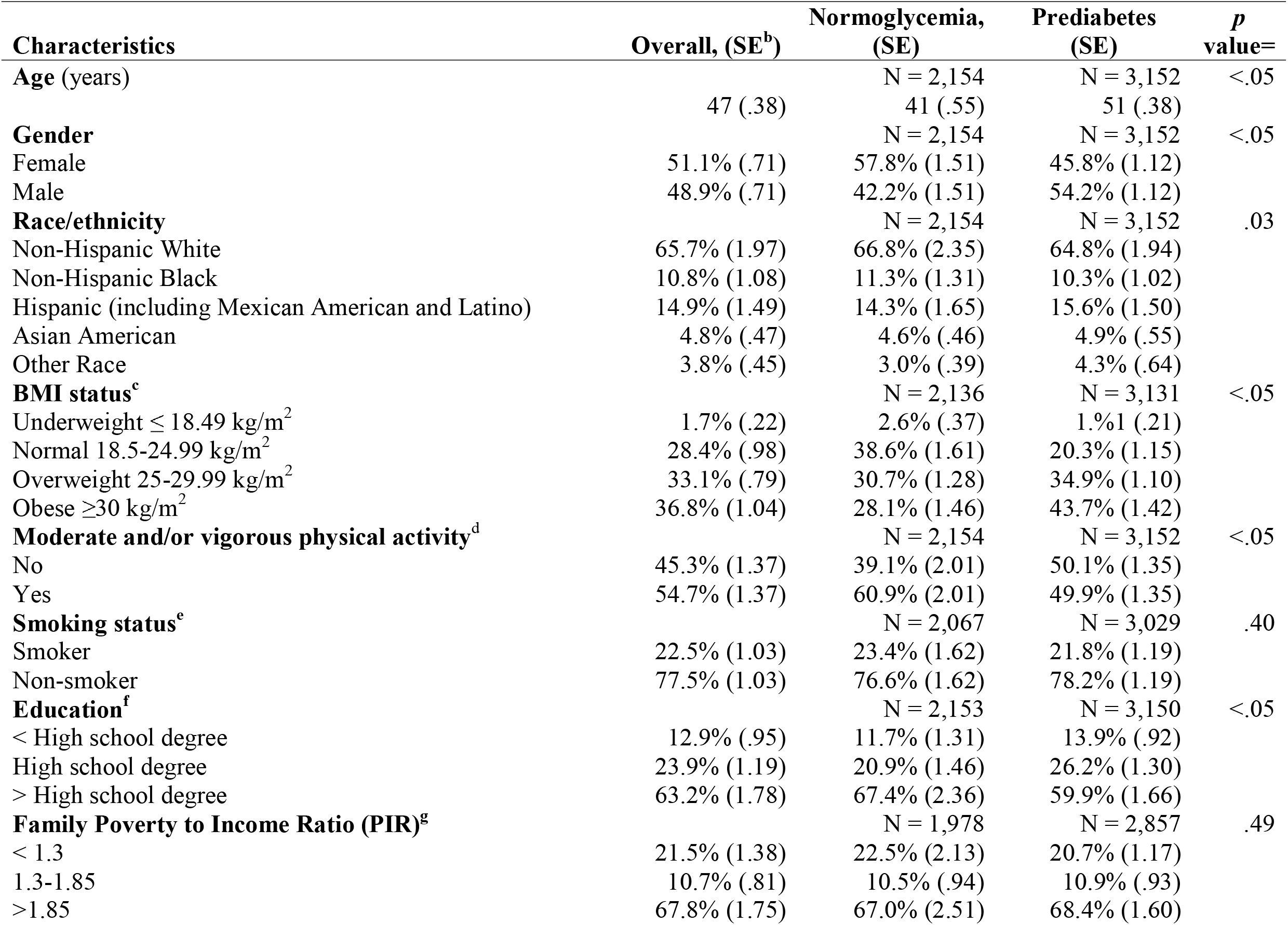
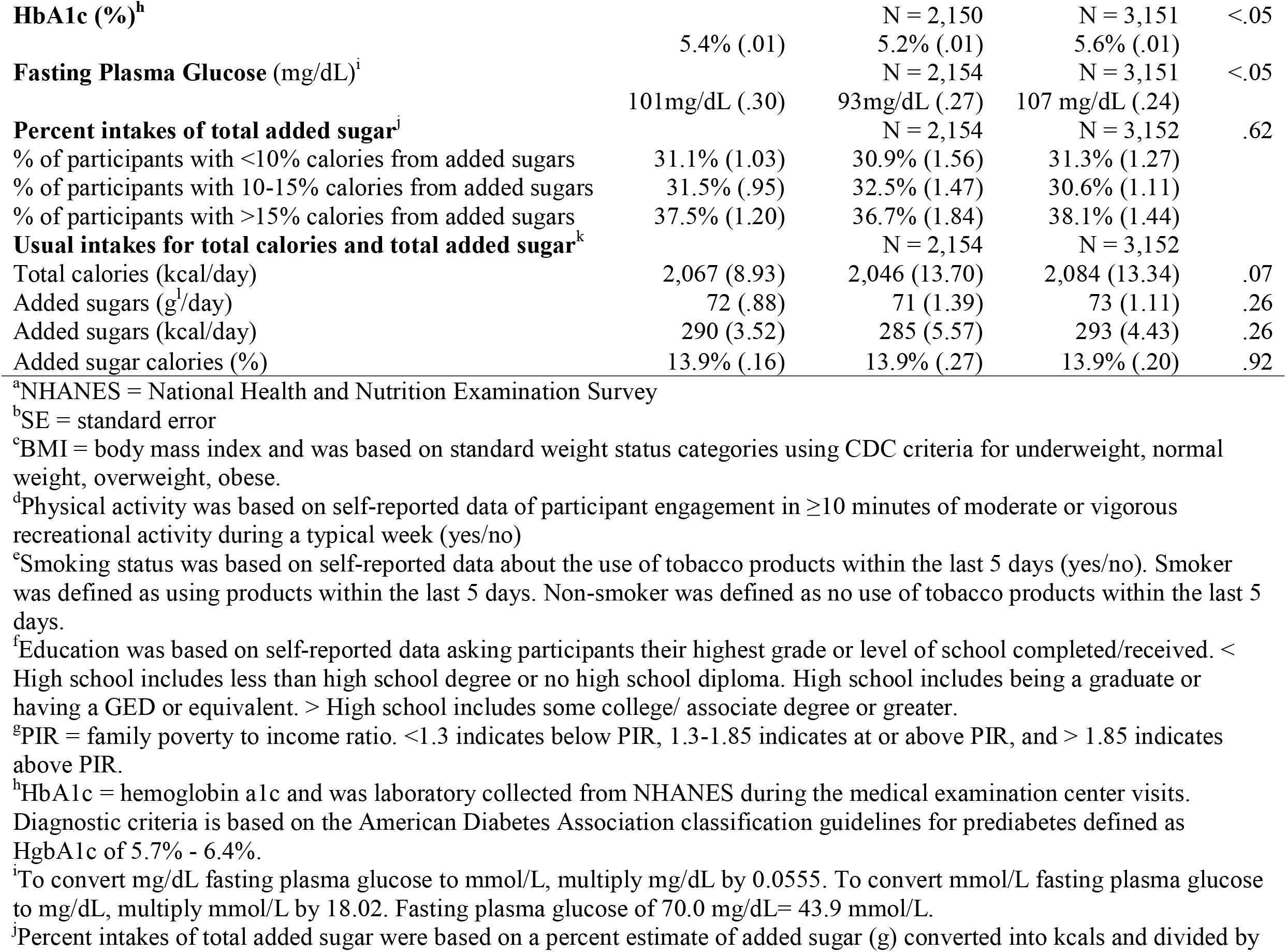
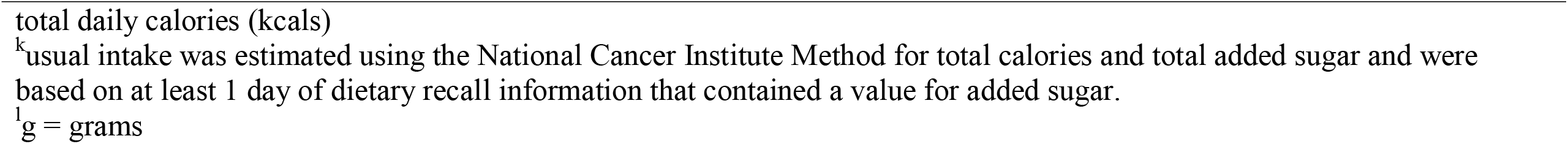
Overall characteristics of adults ≥20 years and by normoglycemia and prediabetes status, the NHANES^a^ 2013-2018

In the overall sample, the average age was 47 years and included more females (51.1%) than males (48.9%) and mostly non-Hispanic White adults (65.7%), followed by Hispanic including Mexican American and Latino (14.9%), non-Hispanic Black (10.8%), Asian American (4.8%), or Other Race (3.8%) adults. The majority of the sample had obesity (36.8%), reported engaging in moderate and/or vigorous physical activity (54.7%), reported being non-smokers (77.5%), had more than a high school degree (63.2%), and reported a family income that represented a PIR >1.85 (67.8%). The average HbA1c was 5.4% and the average FPG was 101 mg/dL. Usual intakes for total calories were 2,067 kcal/day and usual intakes of total added sugar were 72 grams (290 kcal/day).

Comparing between participants (normoglycemia vs prediabetes), those with prediabetes were more likely to be older (51 years), to be male (54.2%), to identify as being of Hispanic including Mexican American (15.6%), Asian American (4.9%), or Other Race (4.3%), have overweight (34.9%) or obesity (43.7%), be less likely to engage in moderate and/or vigorous activity (50.1%), identify as a non-smoker (78.2%), report having a high school (26.2%) or less than high school degree (13.9%) and report a family income that represented a PIR >1.85 (68.4%). Those with prediabetes had an average HbA1c of 5.6% and a FPG of 107 mg/dL compared to an average HbA1c of 5.2% and FPG of 93 mg/dL among adults with normoglycemia. Between the normoglycemia and prediabetes groups, total calories per day and added sugar intake was similar (2,084 vs 2,046 total kcal/day [*p* = .07]; 71 vs. 73 g/day added sugar [*p* = .26]; 285 vs. 293 kcal/day of added sugar [*p* = .26]). Total energy intake from added sugar for both groups was 13.9% (*p* = .92). Percent intakes of added sugar were also similar between groups (Table 1).

### Added Sugar Intake and Prediabetes Risk

#### Total Added Sugar Intakes

Findings from both unadjusted and adjusted models (Table 2) indicated that total added sugar (g/day) intake did not significantly increase the odds of having prediabetes (unadjusted: OR: 1.01, 95% CI: .99 - 1.00, *p* = .26; adjusted: OR: 1.00, 95% CI: .99 - 1.00, *p* = .91). Differences in the odds for having prediabetes were observed for some covariates in the adjusted model. For example, being older, being Hispanic, Asian American or Other Race, and having obesity was associated with a greater odds of having prediabetes, whereas being a non-smoker or having an education beyond a high school degree (relative to no high school degree) was associated with a lower odds of having prediabetes (Table 2).

**Table 2.**
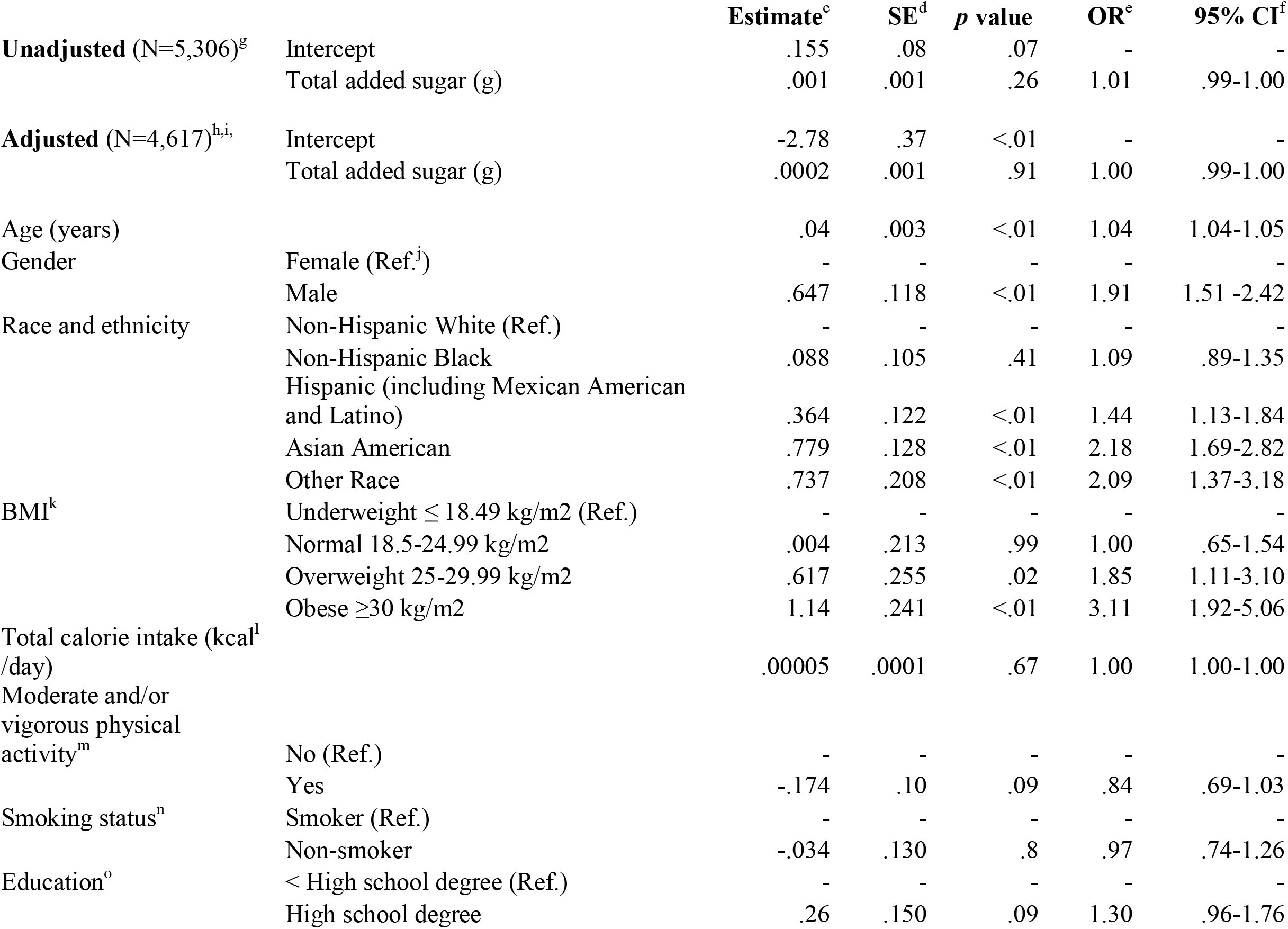
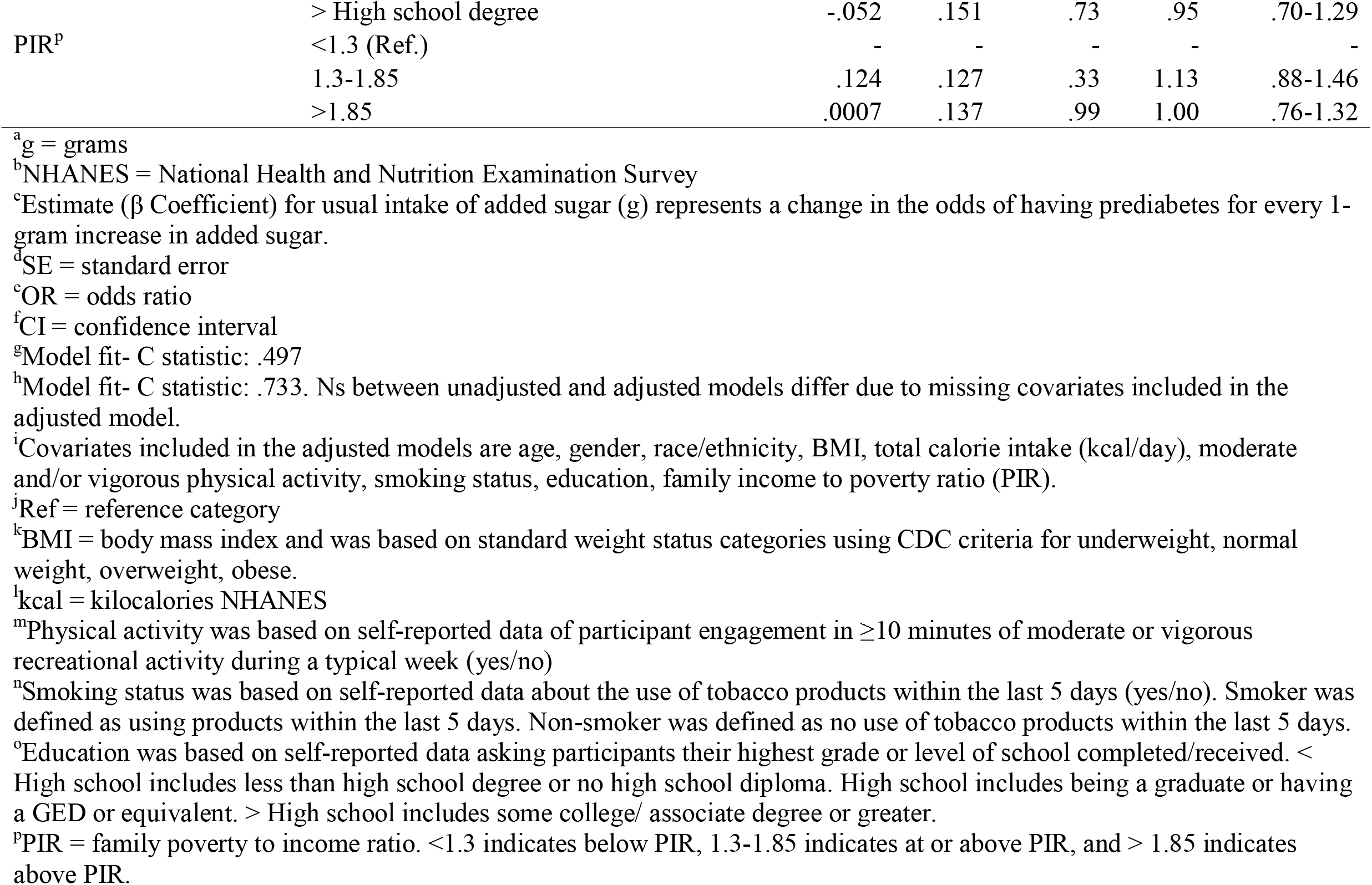
Unadjusted and adjusted odds of prediabetes for total added sugar (g^a^) in U.S. adults ≥20 years with normoglycemia and prediabetes, the NHANES^b^ 2013-2018

Table 3 reports the estimated probability (i.e., risk) for prediabetes at mean (73 g/day) and tertial intakes (43 g/day, 64 g/day, 93 g/day) for total added sugar estimated from unadjusted and adjusted models. In both unadjusted and adjusted models, differences in the estimated risk for prediabetes and total added sugar intake were not statistically significant (unadjusted: *p*=.26; adjusted: *p*=.91). For example, in the unadjusted model, the estimated risk for prediabetes at mean and tertial intakes for total added sugar ranged from 55.8% (mean) and from 55% to 55.6% to 56.4% (tertial). Estimated risk percentages were converted from the ‘estimates’ reported in Table 3 (e.g., .550 equates to 55%). Similarly, in the adjusted models, the estimated risk for prediabetes at mean and tertial intakes for total added sugar ranged from 60.7% (mean) and from 60.6% to 60.6% to 60.8% (tertial) indicating very little difference in estimated risk of prediabetes between varying amounts of total added sugar.

**Table 3.** Estimated risk of prediabetes at mean and tertials of total added sugar (g^a^) in U.S. adults ≥20 years, the NHANES^b^ 2013-2018

|  | **Total Added Sugar** | **Estimate<sup>c</sup>** | **SE<sup>d</sup>** |
| --- | --- | --- | --- |
| **Unadjusted<sup>e</sup>** | Mean (73g) | .558 | .012 |
| Prediabetes (N=3,152) | 1st Q<sup>f</sup> (43g) | .550 | .014 |
|  | Median Q (64g) | .556 | .012 |
|  | 3rd Q (93g) | .564 | .013 |
| **Adjusted<sup>g,h</sup>** | Mean (73g) | .607 | .018 |
| Prediabetes (N=2,735) | 1st Q (43g) | .606 | .020 |
|  | Median Q (64g) | .606 | .018 |
|  | 3rd Q (93g) | .608 | .020 |

<sup>a</sup>g = grams
<sup>b</sup>NHANES = National Health and Nutrition Examination Survey
<sup>c</sup>Estimates represent the risk probability for prediabetes based on mean and tertial intakes of total added sugar (g/day).
<sup>d</sup>SE = standard error
<sup>e</sup>Model fit- C statistic: .497 and p-value = .26
<sup>f</sup>Q = quartiles
<sup>g</sup>Model fit- C statistic: .733 and p-value = .91. Ns between unadjusted and adjusted models differ due to missing covariates included in the adjusted model.
<sup>h</sup>Adjusted for age, gender, race/ethnicity, BMI, total calorie intake (kcal/day), moderate and/or vigorous physical activity, smoking status, education, family poverty to income ratio (PIR).

#### Percent Intakes of Added Sugar

In both unadjusted and adjusted models (Table 4), consumption of different percent intakes of added sugar (<10%, 10-15%, >15%) did not significantly increase the odds of having prediabetes (unadjusted [<10%: (ref); 10-15%: OR: .93, 95% CI: .77 - 1.12, *p* = .44; >15%: OR: 1.03, 95% CI: .82 - 1.28, *p* = .82] and adjusted [<10%: (ref); 10 - 15%: OR: .82, 95% CI: .65 - 1.04, *p* = .09; >15%: OR: .96, 95% CI: .74 - 1.24, *p* = .73]). In the adjusted model, significant differences in the odds for prediabetes were noted for certain covariates including age, race/ethnicity, BMI, smoking status, and education (Table 4). Findings were similar to what was previously reported for the total added sugar adjusted model in Table 2.

**Table 4.**
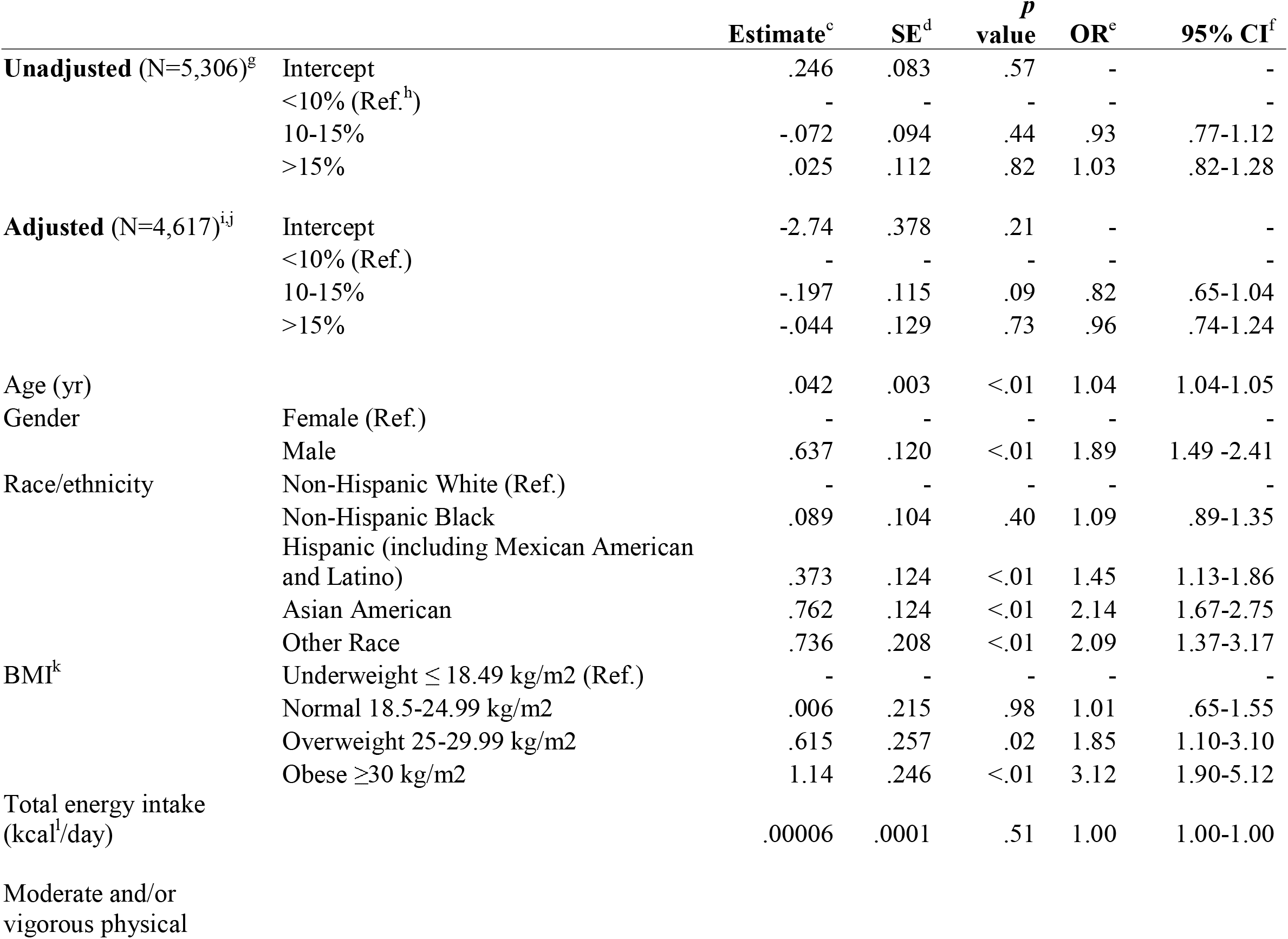
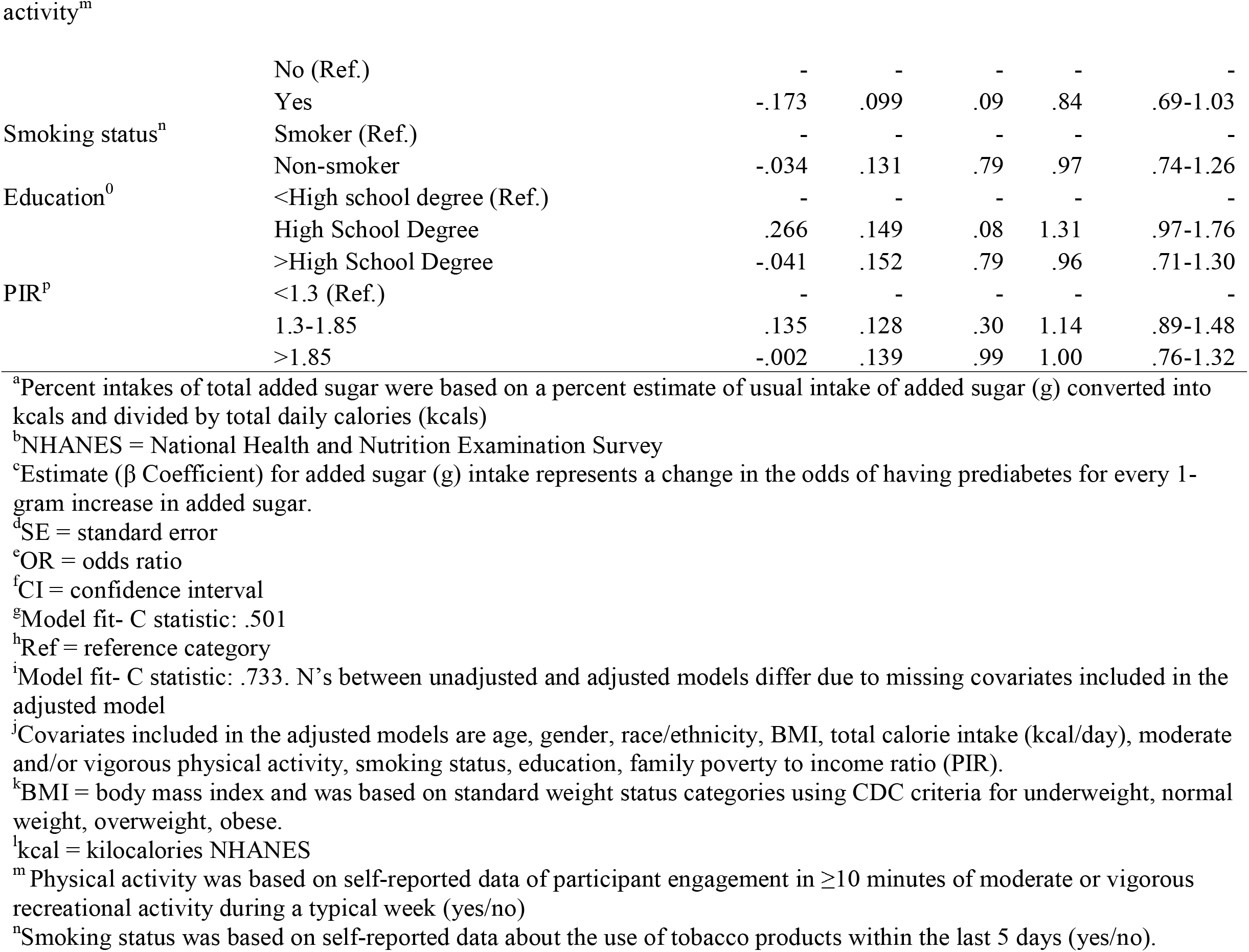
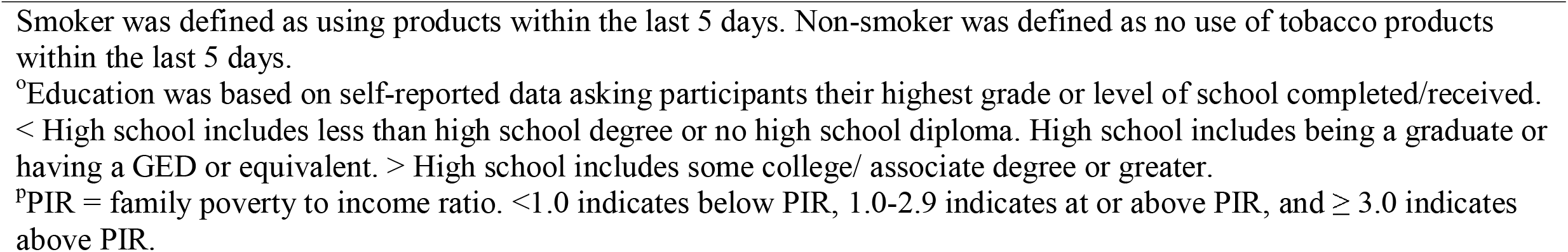
Unadjusted and adjusted odds of prediabetes for percent intakes of total added sugar^a^ in U.S. adults ≥20 years with normoglycemia and prediabetes, the NHANES^b^ 2013-2018

The estimated risk of prediabetes by percent intakes of added sugar (<10%, 10-15%, >15%) was reported in unadjusted and adjusted models (Table 5). Similar to the findings for the total added sugar intake models (Table 3), differences in the estimated risk for prediabetes in the percent intake models were relatively small and not statistically significant (unadjusted: *p* = .51; adjusted: *p* = .22). For example, in the unadjusted model, the estimated risk for prediabetes by percent intakes ranged from 54.3% to 56.1% to 56.7% and in the adjusted model ranged from 57.7% to 61.4% to 62.5% (Table 5).

**Table 5.** Estimated risk of prediabetes for percent intake of total added sugar^a^ in U.S. adults ≥20 years, the NHANES^b^ 2013-2018

|  | **% Added Sugar** | **Estimate<sup>c</sup>** | **SE<sup>d</sup>** |
| --- | --- | --- | --- |
| **Unadjusted<sup>e</sup>** | <10% | .561 | .020 |
| Prediabetes (N=3,152) | 10-15% | .543 | .018 |
|  | >15% | .567 | .019 |
| **Adjusted<sup>f,g</sup>** | <10% | .625 | .021 |
| Prediabetes (N=2,735) | 10-15% | .577 | .029 |
|  | >15% | .614 | .025 |

<sup>a</sup>Percent intakes of total added sugar were based on a percent estimate of usual intake of added sugar (g) converted into kcals and divided by total daily calories (kcals)
<sup>b</sup>NHANES = National Health and Nutrition Examination Survey
<sup>c</sup>Estimate represents the risk probability for prediabetes based on mean and quartile intakes of added sugar in grams per day.
<sup>d</sup>SE = standard error
<sup>e</sup>Model fit- C statistic: .501 and p-value = .51
<sup>f</sup>Model fit- C statistic: .733 and p-value = .22. Ns between unadjusted and adjusted models differ due to missing covariates included in the adjusted model.
<sup>g</sup>Adjusted for age, gender, race/ethnicity, BMI, total calorie intake (kcal/day), moderate and/or vigorous physical activity, smoking status, education, family poverty to income ratio (PIR).

#### Total and Percent Intake of Added Sugar by Race and Ethnicity

Results from the sensitivity analyses (Table 6 and 7) indicated that the association between total and percent intakes of added sugar and risk for prediabetes did not differ by race/ethnicity (Type 3 tests for interaction of race/ethnicity by total added sugar: unadjusted model [*p* = .27]; adjusted model [*p* = .33] and percent intake of added sugar: unadjusted model [*p* = .21]; adjusted model [*p* = .11]). Irrespective of added sugar, it was observed that some racial/ethnic groups had higher odds for prediabetes. In Table 6, adjusted models indicated that the risk for prediabetes was high among those who identified as being Hispanic, Asian American, or Other Race with Asian Americans having the highest risk estimates (69% to 73%). Results were similar in Table 7 with adjusted risk estimates for Asian Americans between 71% to 76%.

**Table 6.**
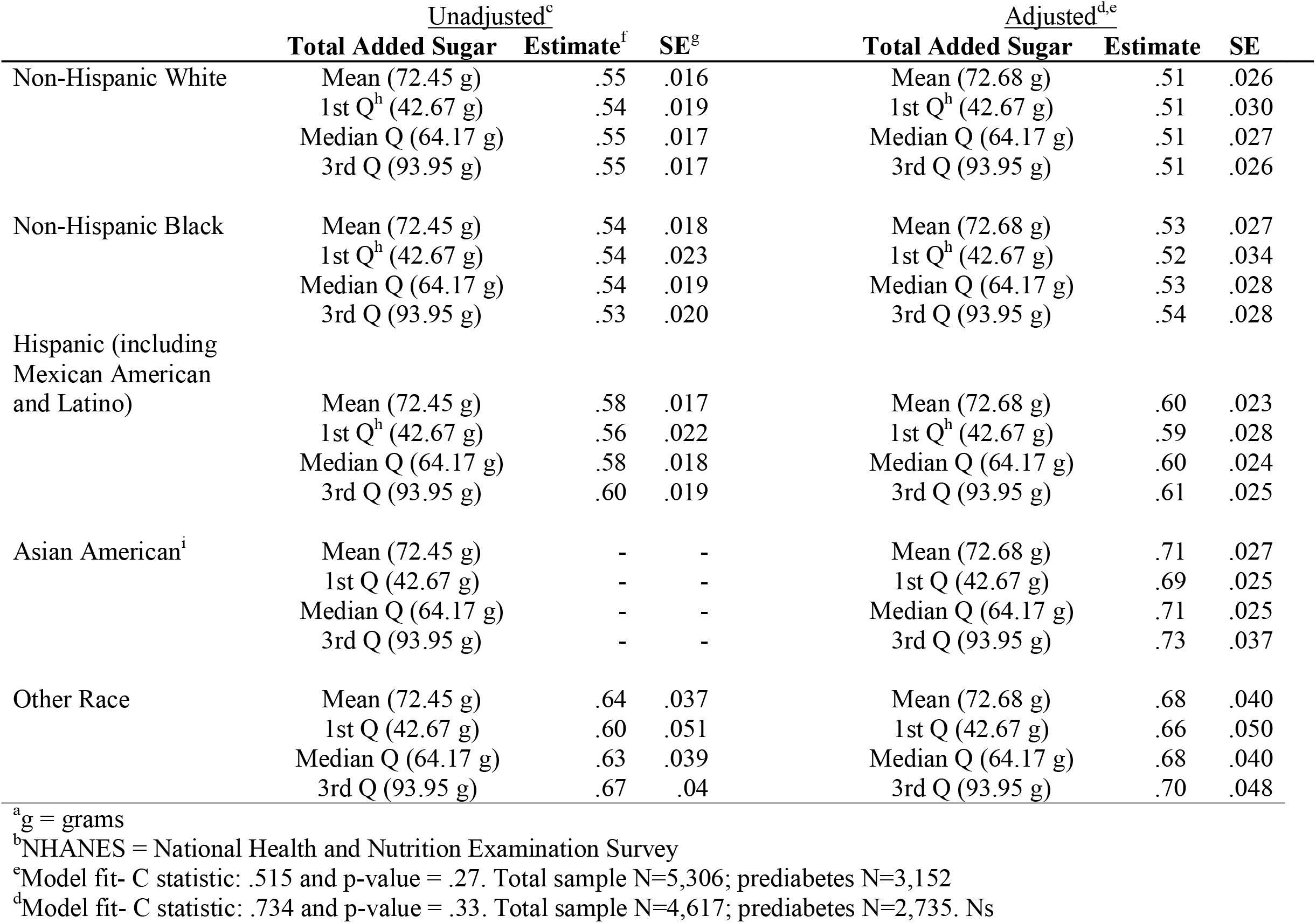
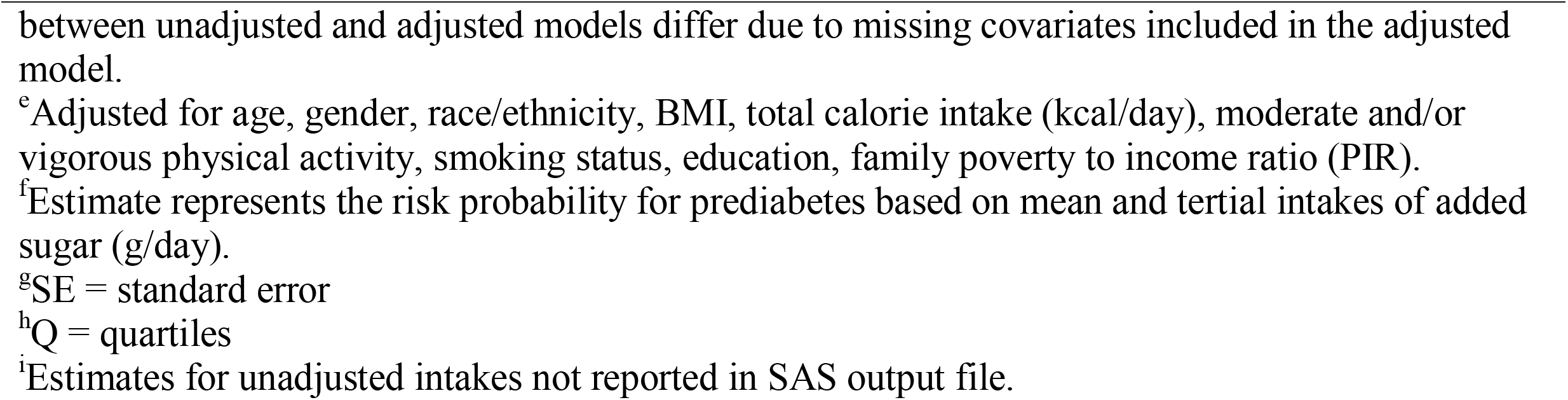
Estimated risk of prediabetes at mean and tertials of total added sugar (g^a^) by race/ethnicity status for U.S. adults ≥ 20 years, the NHANES^b^ 2013-2018

**Table 7.**
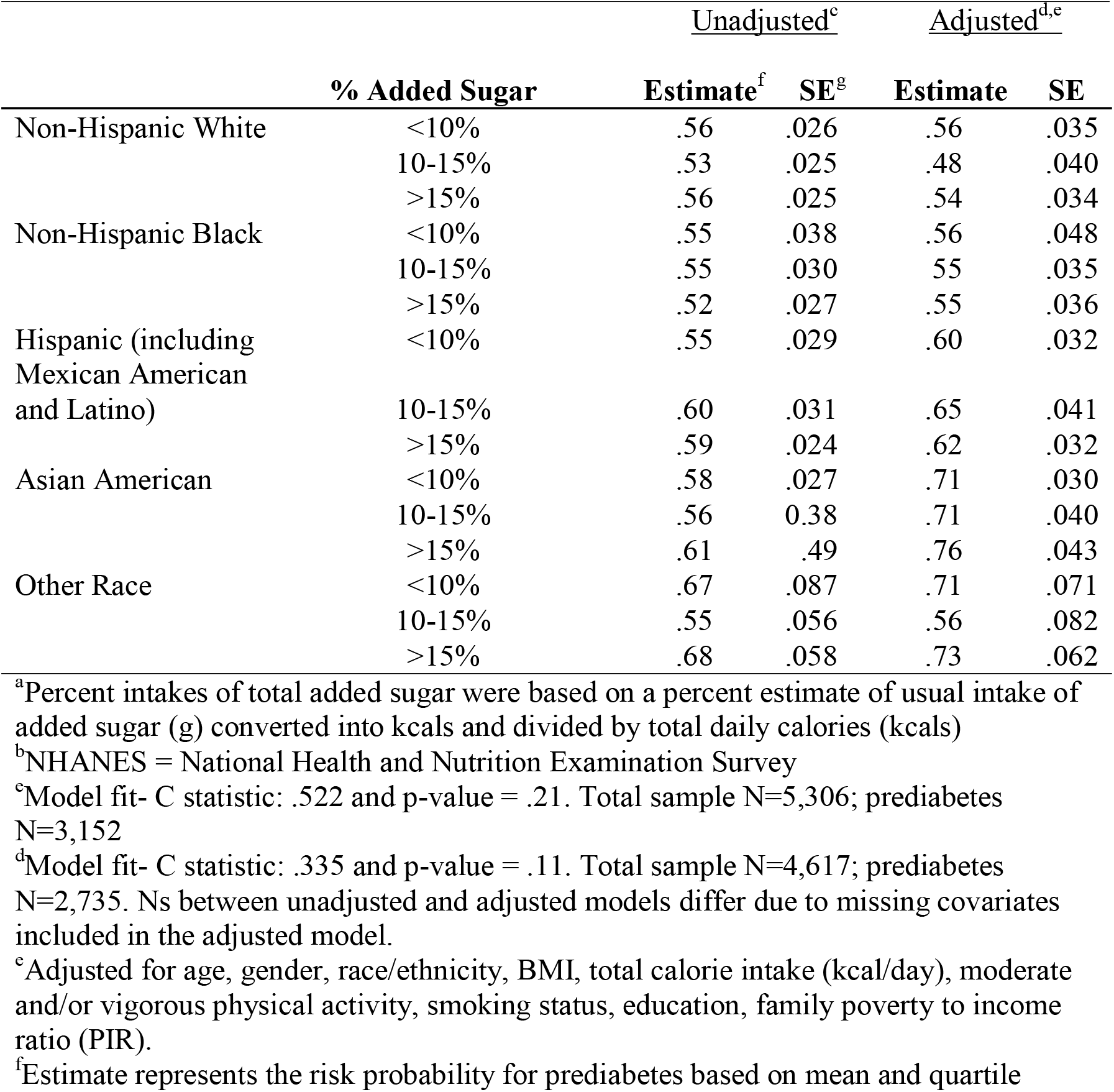
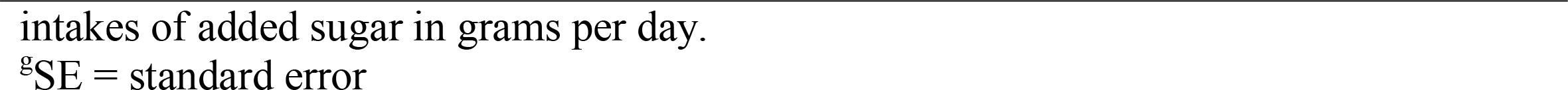
Estimated risk of prediabetes by percent intakes of total added sugar^a^ consumed by race/ethnicity status for U.S. adults ≥ 20 years, the NHANES^b^ 2013-2018

## Discussion

This is the first known study to examine associations between usual intake of *total* added sugar and prediabetes risk in a large nationally representative sample of 5,306 U.S. adults ≥ 20 years. In the study, added sugar accounted for 13.9% of the sample’s total energy intake. Similarly, total energy intake from added sugar in adults with normoglycemia and prediabetes was 13.9%. The findings of this study suggest that even after controlling for total calorie intake, BMI, and pertinent health behaviors/sociodemographic factors, all sources (i.e., total) of added sugar consumed as part of a usual diet, do not appear to significantly increase an individual’s odds for having prediabetes. Average intakes for added sugar in this study exceeded current dietary guideline recommendations and were very close to average intake estimates reported for the total U.S. population (13.9% total energy intake/290 calories per day vs. 13% total energy intake/270 calories per day respectively).^22^

The main hypothesis of this study, that total added sugar consumption would be associated with and increase the risk of prediabetes, was not supported by the findings from this study. Nonetheless, the results are not in complete contrast to what has previously been reported in the literature. Evidence from both observational and experimental findings on this topic have been mixed with many studies suggesting that added sugar increases the risk for prediabetes^24,31-33,36-39^ and fewer studies reporting no such relationship.^71-77^ Yet, differences between study designs (observational vs experimental), characterization of prediabetes risk, inclusion of representative minority groups, and operationalization of added sugar (i.e., proxies such as SSB or fructose-only beverages used to represent total added sugar intake) lend themselves to the inconsistent findings between many studies.

For example, experimental and observational studies have used a wide range of glycemic variables to assess prediabetes risk. In experimental studies, insulin sensitivity has been measured using the hyperinsulinemic clamp,^25,37,73,75,78^ the hepatic insulin sensitivity index,^39^ or a 75 g oral-glucose tolerance test (OGTT)^40^ whereas insulin resistance has been measured via the homeostatic model assessment of insulin resistance (HOMA-IR).^72,74,76,79^ In observational studies, HOMA-IR has commonly been used to assess insulin resistance.^31-33,36^ Interestingly, only a single, prospective cohort study has reported a significant association between added sugar and incident prediabetes measured via FPG and OGTT.^36^ In contrast, prediabetes risk in this study was defined using HbA1c and FPG. Because OGTT was not collected in the 2017-2018 NHANES dataset, it was not used to identify prediabetes. It is possible that including all prediabetes measurements (HbA1c, FPG, and OGTT) may have resulted in a significant association between added sugar and prediabetes risk.

Sample characteristics have also varied widely between studies with experimental studies predominately using a homogenous sample of either male-only^25,37-39,73,74^ or female-only participants^72^ whereas observational studies have mainly included heterogeneous samples.^31-33,36^ Most of the studies failed to consider differences in added sugar intake by race/ethnicity status which limits the generalizability of past findings. In this study, a diverse and heterogeneous sample was included to test whether the associations between added sugar consumption and prediabetes differed by race/ethnicity. The findings of this study do not indicate prediabetes risk differs by race/ethnicity status for total or percent added sugar intakes. However, one notable finding from this study was that the highest risk estimates for prediabetes (irrespective of added sugar intake) was observed among Asian American adults. This is of interest since national estimates indicate that Asian American adults have the lowest incidence of prediabetes in the U.S. compared to other racial/ethnic groups (i.e., 32.8% compared to 35.4% for Hispanic, 36.9% for non-Hispanic Black, and 33.9% for non-Hispanic White adults). Research suggests Asian American adults often have higher rates of prediabetes at lower BMIs (underweight to obese class I).^5^ Participants in this study were predominately overweight or obesity; however, differences in BMI status by race/ethnicity were not estimated. Future studies should specifically include Asian Americans to better characterize their prediabetes risk.

Operationalization of added sugar has also varied widely between studies. For example, a plethora of observational studies have primarily relied on added sugar proxies (e.g., SSB sweetened with HFCS) to approximate total added sugar intake which has been shown to be strongly correlated with risk for prediabetes and T2D.^31-36^ Other studies have used fructose to represent added sugar and have found that greater concentrations of fructose (15% to ≥ 25% of total calories) promote insulin resistance,^37-39^ increase fasting plasma glucose concentrations,^40^ and impair insulin sensitivity.^38-40^ Yet, in the U.S., a typical ad libitum diet consists of different types (e.g., sucrose, glucose, fructose, HFCS) of added sugar that are consumed in both solid and liquid form (e.g., baked goods or beverages like sodas and fruit drinks).^18,22,80^ Aware of this knowledge, the objective of this study was to examine the relationship between *all* (i.e., total) added sugars (consumed as part of a usual diet) and risk for prediabetes among U.S. adults. Because this was a cross-sectional study, causal inferences could not be made to determine why total added sugar intake did not increase the risk for prediabetes in the study sample. With much of the literature pointing to SSBs as a driving factor for diabetes risk,^31-36^ it may be that liquid sources (e.g., SSBs) of added sugar compared to solid sources (e.g., baked goods, confectionaries) pose a greater risk for prediabetes. This was beyond this scope of this study; however, a few studies have found that added sugar consumed from liquids are responsible for impaired glucose homeostasis and insulin resistance in children (8-10 years)^81^ and positively correlated with HOMA-IR in adults whereas similar correlations have not observed for solid food consumption.^82^ Experimental research is needed to explore these differences in the context of prediabetes.

Lastly, it should be noted that the findings from this study are not in complete contrast to what has been previously reported in the literature. In a study by Lowndes et al.,^76^ added sugar represented ∼18% of the sample’s total energy intake and did not impair fasting glucose concentrations in adults without diabetes (pre- or type 2). In comparison, consumption of added sugar in this study was slightly lower than what was used in the study by Lowndes et al. It is possible that average intakes of ∼14% to ∼18% added sugar may not pose a significant risk for prediabetes in adults. However, experimental studies assessing varying intakes of total added sugar as part of an ad libitum diet are needed since Lowndes et al. supplemented sucrose to participants in liquid form (via unsweetened milk), which as previously mentioned, may have differing effects on risk for prediabetes.^81,82^

### Strengths and Limitations

This study has some major strengths. This is the first, known study to assess the associations between total and percent intakes of added sugar and risk for prediabetes in a nationally representative sample of U.S. adults. Differences by race and ethnicity status were also examined to improve the generalizability of the study results. Additionally, the NCI method was used to estimate usual individual intakes for added sugar for use in a disease model which helps account for between- and within-person variation in intake.^66,83^

This study also has limitations. First, the cross-sectional nature of this study did not allow for the assessment of causal or temporal inferences between added sugar and risk for prediabetes. Second, self-reported 24-hour dietary recalls were used to estimate usual intake of added sugars and total calories which may be subject to under- or overreporting due to concerns of social desirability.^84^ However, use of the AMPM method, which has been found to accurately estimate usual nutrient intake, may have reduced this concern.^85,86^ Third, HbA1c is reported to have a lower sensitivity at cut-points of 5.7-6.4% and is associated with greater diagnostic inaccuracy in the presence of certain medical conditions that increase red blood cell turnover (e.g., sickle cell disease, pregnancy, erythropoietin therapy)^1^ and are not reported in NHANES. Differences by race and ethnicity status have also been reported with HbA1c levels registering higher in non-Hispanic Black adults compared to non-Hispanic White adults who had similar fasting glucose levels.^1^ It is possible some individuals were incorrectly classified as having prediabetes (or T2D); however, including both FPG and HbA1c likely improved identification of prediabetes among the study sample.^87^

## Conclusions

This study found that added sugar averaged 13.9% of the sample’s total energy intake and was the same for adults with normoglycemia and prediabetes. Total and percent intakes of added sugar did not increase the risk for prediabetes in this nationally representative study of U.S. adults ≥20 years, including no significant differences in risk by race/ethnicity status. As this topic continues to evolve, additional experimental studies are needed to determine if there are any direct effects from consuming *total* added sugar, as part of a usual diet, on prediabetes risk. As rates of prediabetes and T2D continue to rise, evidence-based research should determine what role added sugar plays in diabetes management and prevention to help advance the field of precision health and nutrition.

## Data Availability

All data produced in the present study are available upon reasonable request to the authors

## ACKNOWLEDGEMENTS

The authors sincerely thank Dr. Greg Pavela for his conceptual, methodological, and statistical expertise required for completion of this manuscript and for his critical manuscript review. (I have received permission from those named in the acknowledgement.)

